# Multifaceted Brain Age Measures Reveal Premature Brain Aging and Associations with Clinical Manifestations in Schizophrenia

**DOI:** 10.1101/2020.11.09.20228064

**Authors:** Chang-Le Chen, Tzung-Jeng Hwang, Yu-Hung Tung, Li-Ying Yang, Yung-Chin Hsu, Chih-Min Liu, Hai-Gwo Hwu, Yi-Tin Lin, Ming-Hsien Hsieh, Chen-Chung Liu, Yi-Ling Chien, Wen-Yih Isaac Tseng

## Abstract

Schizophrenia is a mental disorder with extensive alterations of cerebral gray matter (GM) and white matter (WM) and is known to have advanced brain aging. However, how the structural alterations contribute to brain aging and how brain aging is related to clinical manifestations remain unclear. Here, we estimated the bias-free multifaceted brain age measures in patients with schizophrenia (N=147) using structural and diffusion magnetic resonance imaging data. We calculated feature importance to estimate regional contributions to advanced brain aging in schizophrenia. Furthermore, regression analyses were conducted to test the associations of brain age with illness duration, onset age, symptom severity, and intelligence quotient. The patients with schizophrenia manifested significantly old-appearing brain age (*P*<.001) in both GM and WM compared with the healthy norm. The GM and WM structures contributing to the advanced brain aging were mostly located in the frontal and temporal lobes. Among the features, the GM volume and mean diffusivity of WM were most sensitive to the neuropathological changes in schizophrenia. The WM brain age index was associated with a negative symptom score (*P*=.006), and the WM and multimodal brain age indices demonstrated negative associations with the intelligence quotient (*P*=.037; *P*=.040, respectively). Moreover, brain age exhibited associations with the onset age (*P*=.006) but no associations with the illness duration, which may support the early-hit non-progression hypothesis. In conclusion, our study reveals the structural underpinnings of premature brain aging in schizophrenia and its clinical significance. The brain age measures might be a potentially informative biomarker for stratification and prognostication of patients with schizophrenia.

## 1 Introduction

A growing body of evidence has demonstrated that schizophrenia is a psychiatric disorder with neurobiological alterations involved in both neurodevelopmental and neurodegenerative processes^1-3^ that manifest various impairments in brain structure and function^4-6^. Neuroimaging studies have reported pronounced gray matter (GM) volume loss throughout the brain^7,8^ and reduction in cortical thickness primarily in the frontal and temporal areas^9^. These changes resemble the changes observed in the aging process^10^. Moreover, diffusion magnetic resonance imaging (MRI) constantly reported altered integrity of white matter (WM) in schizophrenia^11^ which reflects the disconnection between cortical areas and may lead to cognitive impairments^12^. One study reported that WM integrity, as measured by fractional anisotropy, was reduced in younger patients with schizophrenia, and the reduction pattern was similar to that in older healthy controls^13^, suggesting a premature reduction of WM integrity in schizophrenia. These findings demonstrate an older biological status of the brain in patients and suggest premature brain aging in schizophrenia^14,15^.

A neuroimaging-based brain age paradigm has been developed as an imaging biomarker to investigate aberrant brain aging, which occurs in numerous neurological diseases and psychiatric disorders^14,16-18^. This approach enables precise and individualized quantification of the extent of brain aging. To estimate the brain age index, numerous brain scans are acquired from cognitively healthy individuals, and these brain scans are employed as a reference cohort to create and define a brain age prediction model. Through modern machine learning and deep learning techniques, brain scans can be transformed from high-dimensional neuroimaging features into a concise brain age marker by learning the complex aging pattern in biomedical images. Consequently, the established brain age prediction model can predict the brain age of other individuals. The predicted age difference (PAD), which is the difference between an individual’s brain age and chronological age, quantitatively indicates deviation from the defined healthy aging trajectory^19^. Depending on the neuroimaging data modality, the derived PAD highlights different aspects of brain aging.

Brain age measures are capable of revealing the underlying mechanism of brain aging in schizophrenia. GM-based brain age measures have indicated apparent brain aging in patients with schizophrenia compared with healthy controls, for those in early stages and those with chronic illness^14,20-22^. Furthermore, several studies have indicated that the effect of apparent brain aging becomes more prominent over time, especially within the short-term period after disease onset, suggesting that accelerated brain aging occurs in patients with schizophrenia^14,22^. However, a study reported that patient’s brain age did not progress for the remainder of life, implying that the aging induced by the disease was not accelerated^23^. The conflicting reports indicate that the hypothesis of accelerated brain aging in schizophrenia is controversial.

Although these pioneering studies demonstrated the applications of brain age paradigms in schizophrenia, several limitations remain. First, most brain age research has only adopted the single imaging modality, such as GM volumetric features, to estimate the PAD. Few studies have addressed the brain aging in schizophrenia with other imaging features such as WM integrity and multimodal features. Multidimensional investigations of brain structure alterations in patients with schizophrenia are still lacking. Moreover, regular PAD, which is the most common measure employed in brain age studies, has an intrinsic statistical bias when estimated during the model training phase; the PAD is significantly correlated with chronological age in the model training set^24^. This bias makes PAD dependent on chronological age. To our knowledge, no study of brain age in schizophrenia has directly addressed this problem. Furthermore, although machine-learning–based brain age predictions provide an advanced approach for quantitatively estimating the degree of brain aging, the characteristics of the unexplainable “black box” inside the machine learning algorithms hinder the interpretation of feature importance for neurobiological inference.

To investigate the multifaceted biological age in schizophrenia and address these limitations, we constructed three types of brain age models based on the neuroimaging features of the GM, WM, and their combination (i.e., multimodality). We then estimated the bias-free brain age indices of patients with schizophrenia. We hypothesized that these brain age indices may reflect different aspects of the advanced brain aging in the patients with schizophrenia. Additionally, the clinical relevance of brain age indices in schizophrenia was investigated; we postulated that brain age indices may exhibit statistical associations with various clinical factors, such as illness duration, onset age, symptom severity, and general cognition (e.g., full-scale intelligence quotient, FSIQ). Moreover, we established a series of region-of-interest (ROI)-based normative models^25^ by using healthy individuals to define the norm of imaging measures and then quantify the extent of structural impairments in schizophrenia against the healthy norm. This approach enables quantification of certain impaired brain regions in the patients with schizophrenia. We then evaluated multivariate correlation between the structural deviance of the patients with schizophrenia and their brain age indices, estimating the contribution of each structural feature. Using this framework, we aimed to delineate a brain network that explains the advanced brain aging in schizophrenia.

## 2 Materials and Methods

### 2.1 Participants

Patients with schizophrenia (N = 147; mean age = 31.1; standard deviation [SD] = 8.3; range 16– 62; sex: 46.3% men) were consecutively recruited from the outpatient clinic of the Department of Psychiatry of National Taiwan University Hospital (NTUH) from 2010 to 2017. Patients recruited before 2014 were diagnosed of schizophrenia based on the Diagnostic and Statistical Manual of Mental Disorders-4 (DSM-4) criteria, and their symptoms and clinical presentations satisfied the DSM-5 criteria after rediagnosis. Patients recruited after 2014 were diagnosed using the DSM-5 criteria. Diagnoses of schizophrenia were made after comprehensive chart review and personal interviews performed by experienced psychiatrists. Patients with schizoaffective disorder, bipolar disorder, substance abuse, intellectual disability, major systemic disease, or neurological diseases were excluded. The symptoms at initial recruitment were assessed by the senior psychiatrists from the Department of Psychiatry of the NTUH by using the Positive and Negative Syndrome Scale (PANSS), and the FSIQ were measured by using the Wechsler Adult Intelligence Scale—Third Edition^26,27^. All participants provided written informed consent, and the Institutional Review Board of NTUH approved the study.

Brain images of 482 cognitively normal individuals (mean age = 36.9, SD = 19.1, range = 14– 92; sex: 53.1% women) obtained from the NTUH MRI database, including T1-weighted images and diffusion spectrum imaging (DSI) data sets, were used as the training set to develop brain age prediction models. Another independent set of 70 cognitively normal individuals (mean age = 36.8, SD = 19.9, range = 14–83; sex: 52.2% women) from the database was used to assess the reproducibility of the brain age models. All 552 cognitively normal participants had no history of neurologic or psychiatric illness. Detailed information on the recruitment criteria for cognitively normal individuals is provided in Supplementary Material S1.1. All training and test sets were anonymized.

### 2.2 MRI Image Acquisition

All brain images, including the training and test sets and those from patients with schizophrenia, were acquired using the same 3-Tesla MRI scanner (Tim Trio; Siemens, Erlangen, Germany) with a 32-channel phased-array head coil. High-resolution T1-weighted imaging was performed using a three-dimensional (3D) magnetization-prepared rapid gradient-echo sequence with the isotropic spatial resolution in 1 mm^3^. DSI, which is designed to capture the microstructural integrity of WM, was performed using a pulsed-gradient spin-echo diffusion echo-planar imaging sequence with a twice-refocused balanced echo^28^; the imaging parameters were *b*_max_ = 4000 s/mm^2^ and in-plane spatial resolution = 2.5 × 2.5 mm^2^. The diffusion-encoding acquisition scheme followed the framework of DSI^29^, which comprised 102 diffusion-encoding gradients corresponding to the Cartesian grids in the half-sphere of the 3D diffusion-encoding space (*q*-space)^30^. Each MRI scan included T1-weighted imaging. DSI was completed within 20 minutes. Detailed information on the imaging parameters is provided in Supplementary Material S2.1.

### 2.3 Image Analysis

Before image data analysis was performed, all T1-weighted images and DSI data sets underwent quality assurance procedures, which are detailed in Supplementary Information S2.2. To extract GM features from the T1-weighted images, voxel-based morphometry and surface-based morphometry were performed using the Computational Anatomy Toolbox (CAT12)^31^, which is an extension of Statistical Parametric Mapping 12^32^ (Figure 1A). Voxel-based morphometry was applied to estimate voxel-wise regional volume features according to the LONI probabilistic brain atlas, which contains 56 ROIs^33^. Surface-based morphometry was employed to measure cortical thickness through projection-based thickness estimation^34^. The estimated thickness features were sampled according to the 68 cortical ROIs included in the Desikan–Killiany cortical atlas ^35^. The detailed information of the image processing is provided in Supplementary Information S2.3. Briefly, a total of 56 volumetric features and 68 cortical thickness features were obtained and used to estimate the GM-based brain age.

**Figure 1.**
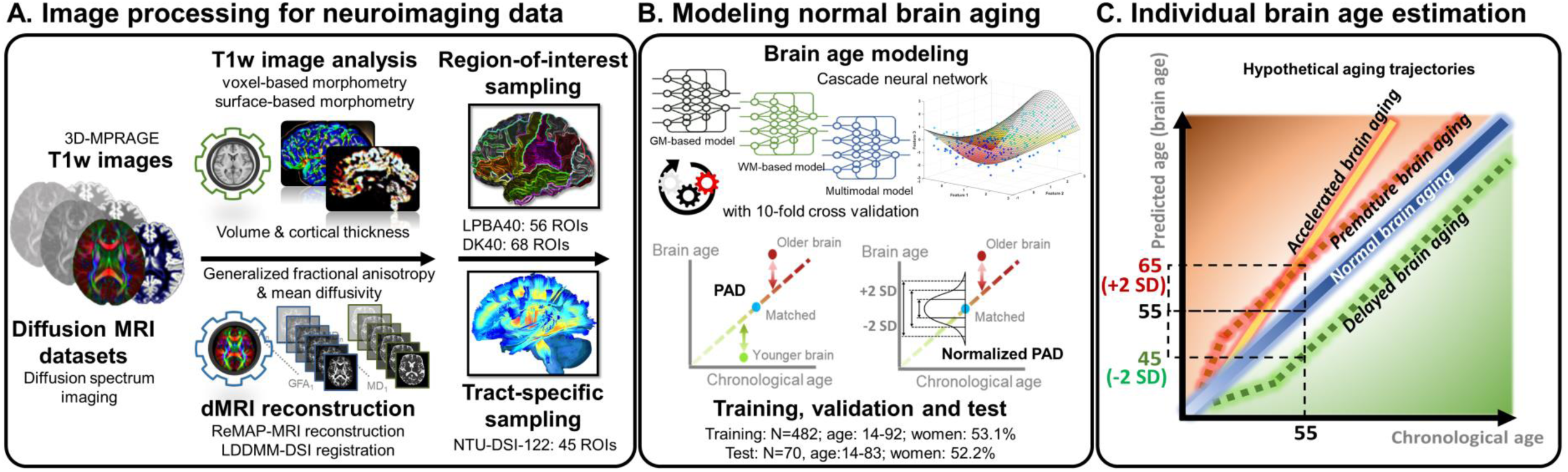
Analytic pipeline and conceptual explanation of brain age. Subplot A illustrates the imaging processing for the T1-weighted images and diffusion spectrum imaging datasets. Subplot B represents the brain age models established using the data sampled from a normal population. Subplot C demonstrates the hypothetical aging trajectories corresponding to the individual brain age inference.

WM features were extracted from DSI data sets by using an in-house automatic analytic pipeline to transform DSI data into tract-specific features^36^ (Figure 1A). The analytic algorithm is detailed in Supplementary Information S2.3. In brief, DSI data were reconstructed into structure-related diffusion indices (i.e., generalized fractional anisotropy [GFA] and mean diffusivity [MD]) by using the regularized version of the mean apparent propagator (MAP)-MRI algorithm^37,38^. A two-step registration process with an advanced diffusion MRI-specific registration algorithm^39^ was employed to minimize the registration bias arising from cross-lifespan data variation. Finally, the predefined tract bundle coordinates on a standard template were projected according to the transformation map obtained from the registration process onto individuals’ diffusion index maps to sample tract-specific features. The pipeline produced 45 tract features for each index from each participant. Consequently, 45 GFA and 45 MD features were obtained to estimate the WM-based brain age. The parcellation of GM and WM ROIs is detailed in Supplementary eTable.

### 2.4 Brain Age Modeling

The GM, WM, and multimodal brain age prediction models were established using the training set’s neuroimaging features to predict brain age based on GM, WM, and both GM and WM features, respectively (Figure 1B). Sex was also included as a predictor. The architecture of the brain age model was designed to have a 12-layer feedforward cascade neural network^40^. A 10-fold cross-validation procedure was performed to estimate the performance of the brain age models in the training phase. An independent test set was then used to evaluate the reproducibility of the brain age models. The detailed description of brain age modeling and results of model performance are provided in Supplementary Information S1.2 and S3, respectively.

After model performance was determined, the correction models for minimizing age-related bias were constructed for each brain age model by using the training set. Age-related bias refers to the statistical bias of PAD (i.e. predicted age − chronological age, used to represent the degree of aging; Figure 1C), which was significantly correlated with chronological age in the training set^24^. In practice, Gaussian process regression (GPR)^41^ was used to obtain regression model estimates for the training set; the independent variables were age and sex, and the dependent variable was the predicted age. The mean and SD of the training sample’s predicted age for a certain age and sex were estimated from the GPR models, and a new individual’s predicted age was standardized into a value resembling a Z-score on the basis of the derived mean and SD of predicted age of the individual’s peers. The Z-score– like value was termed normalized PAD (nPAD). The nPAD value is free of age-related bias and has biological meaning similar to that of the original PAD, with higher values indicating an older brain. This normalization procedure is in accord with the notion of a normative model^42^. Under the normalization, the individual’s predicted age is compared with the individual’s peers’ mean predicted age rather than the individual’s chronological age.

### 2.5 Normative Modeling for Structural Features

ROI-based normative models were also established for each neuroimaging feature by using the training set. The purpose of normative models was to define a statistical norm for each structural feature based on a cognitively normal population-based cohort given certain age and sex^42^. This method enabled the quantification of an individual’s structural deviance compared with the norm. This deviance, which was equivalent to a standardized score (Z-score), served as a measure of the structural integrity of brain regions. A normative model was built for each structural feature (i.e., each GM and WM feature) by using the training set to estimate the mean and SD of model functions with GPR approach. The independent variables were age and sex, and the dependent variable was the structural feature. After ROI-based normative modeling was performed, these normative models were applied to patients with schizophrenia to calculate the Z-score for each brain region. These estimated Z-scores were further used to calculate regional contributions to brain aging in patients with schizophrenia.

### 2.6 Statistical Analysis

Three analyses were performed in the study to test the hypotheses. The first analysis was the comparison of nPAD in schizophrenia with respect to cognitively normal individuals. The nPAD scores derived from GM, WM, and multimodal models were compared with the population mean of cognitively normal individuals, which should be zero, by using one-sample *t* tests. In addition, paired t tests and Pearson’s correlation coefficients were respectively employed to examine the differences and correlation between GM and WM nPAD scores in patients with schizophrenia.

A multiple linear regression analysis was performed to assess the relationship between the nPAD scores (as dependent variables) and clinical manifestations (as independent variables). The clinical manifestations consisted of symptom scores (i.e., PANSS positive, negative, and general scores) and clinical factors (i.e., duration of illness, onset age, and antipsychotic dosage). The multiple comparison problem was addressed by Benjamini-Hochberg correction^43^. In addition, the relationship between brain age indices and general cognition in patients with schizophrenia was assessed by multiple regression analysis associating the nPAD scores (as independent variables) with the FSIQ (as the dependent variable) in the patient group. Age, sex, and years of education were controlled.

The final analysis was to calculate feature importance and identify the regions that contributed most to advanced aging in schizophrenia. The analysis consisted of two steps. First, we calculated patients’ Z-score profiles of structural features using ROI-based normative models. Z-score values indicate the magnitude of structural impairment deviated from the normal means. Second, we tested the correlations between patients’ Z-score values and their nPAD scores using canonical correlation analysis (CCA)^44^. The coefficients of the brain regions in CCA were normalized into 0 to 1. The normalized coefficients represented the weights of the contribution to the nPAD scores, thus reflecting the feature importance of advanced aging in individuals with schizophrenia.

## 3 Results

### 3.1 Comparisons of normalized predicted age difference (nPAD) in schizophrenia

A total of 147 patients with schizophrenia were analyzed in the study. The mean (SD) of duration of illness and age of disease onset were 7.5 (7.0) years and 23.4 (6.9) years, respectively. The PANSS scores were 13.1 (5.1) in positive symptom, 15.8 (7.2) in negative symptom, and 28.2 (8.4) in general symptom. The medication dose was 312.8 (269.8) chlorpromazine-equivalent units at the moment of subject recruitment and FSIQ was 93.8 (12.9) units.

The performance of brain age modeling and nPAD are provided in Supplementary Information S3. After confirming the model performance and verifying the unbiased estimation of nPAD, the nPAD scores of the patients with schizophrenia were estimated for statistical analyses. The results of a mass one-sample *t* test revealed a significant difference in all the nPAD scores of the schizophrenia group compared with the healthy norm (nPAD-GM: 1.03 (1.82), *t*(146) = 6.89, *p* < 0.001; nPAD-WM: 0.84 (1.83), *t*(146) = 5.59, *p* < 0.001; nPAD-multimodal: 1.36 (1.92), *t*(146) = 8.37, *p* < 0.001; adjusted by Benjamini-Hochberg correction) (Figure 2). No significant differences were observed in the nPAD scores of the test set compared with the healthy norm (nPAD-GM: −0.1 (1.40), *t*(69) = −0.60, *p* = 0.553; nPAD-WM: 0.00 (1.06), *t*(69) = −0.03, *p* = 0.977; nPAD-multimodal: −0.07 (1.30), *t*(69) = −0.43, *p* = 0.667). We also determined the original PAD scores, PAD-GM: 5.56 (8.74) years; PAD-WM: 4.00 (9.40) years; PAD-multimodal: 6.13 (8.36) years, which were comparable to the scores reported in the literature. Paired t test showed that there was no significant difference between nPAD-GM and nPAD-WM (*t*(146) = 1.03, *p* = 0.304), but a significant correlation was identified between these two indices (rho = 0.264, *p* = 0.001).

**Figure 2.**
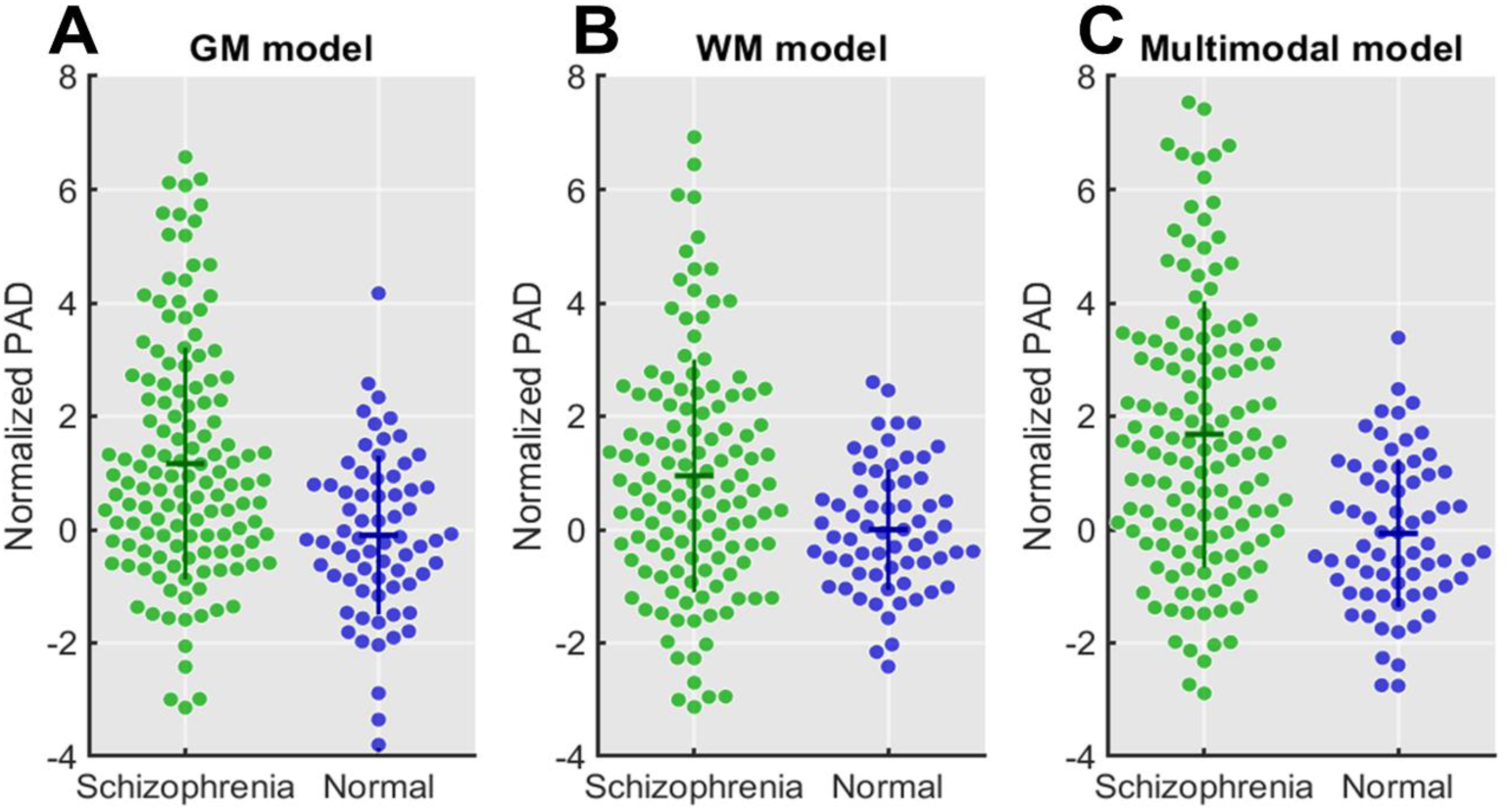
The beeswarm plot of normalized predicted age difference (NPAD) in Schizophrenia and the Normal (test set) based on different brain age models.

### 3.2 Regression analysis of nPAD with symptom scores and clinical factors

Before the regression analysis, patients with clinical factors and symptom scores exceeding SD = 2 were excluded (N = 23) to minimize the result bias caused by outliers. In the regression model of nPAD with symptom scores (Table 1), only negative symptoms were significantly associated with the nPAD-WM (estimated beta = 0.103, *p* = 0.006) and nPAD-multimodal scores (estimated beta = 0.110, *p* = 0.026). In the regression model of nPAD with clinical factors, the age of disease onset exhibited a significant negative correlation with nPAD-WM (estimated beta = −0.107, *p* = 0.006), whereas the duration of illness and antipsychotic dose did not display any significant association with any of the nPAD scores.

**Table 1.**
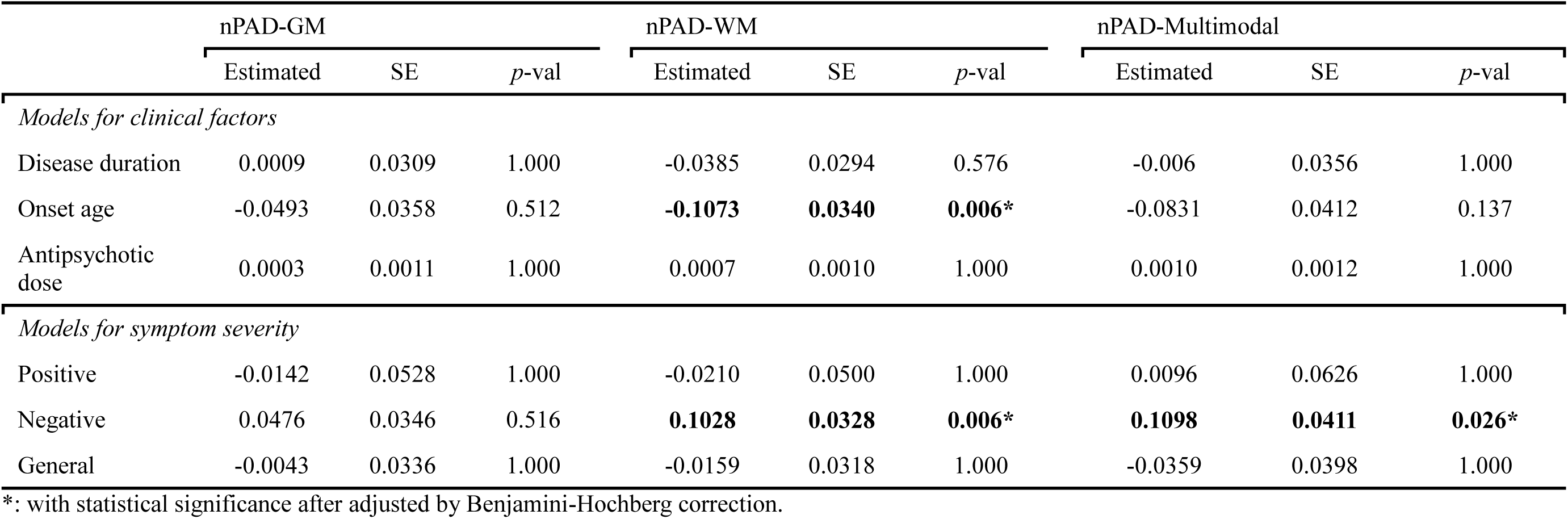
Regression models of nPAD with clinical factors and symptom scores

|  | nPAD-GM |  |  | nPAD-WM |  |  | nPAD-Multimodal |  |  |
| --- | --- | --- | --- | --- | --- | --- | --- | --- | --- |
|  | Estimated | SE | *p*-val | Estimated | SE | *p*-val | Estimated | SE | *p*-val |
| *Models for clinical factors* |  |  |  |  |  |  |  |  |  |
| Disease duration | 0.0009 | 0.0309 | 1.000 | -0.0385 | 0.0294 | 0.576 | -0.006 | 0.0356 | 1.000 |
| Onset age | -0.0493 | 0.0358 | 0.512 | **-0.1073** | **0.0340** | **0.006\*** | -0.0831 | 0.0412 | 0.137 |
| Antipsychotic dose | 0.0003 | 0.0011 | 1.000 | 0.0007 | 0.0010 | 1.000 | 0.0010 | 0.0012 | 1.000 |
| *Models for symptom severity* |  |  |  |  |  |  |  |  |  |
| Positive | -0.0142 | 0.0528 | 1.000 | -0.0210 | 0.0500 | 1.000 | 0.0096 | 0.0626 | 1.000 |
| Negative | 0.0476 | 0.0346 | 0.516 | **0.1028** | **0.0328** | **0.006\*** | **0.1098** | **0.0411** | **0.026\*** |
| General | -0.0043 | 0.0336 | 1.000 | -0.0159 | 0.0318 | 1.000 | -0.0359 | 0.0398 | 1.000 |

\*: with statistical significance after adjusted by Benjamini-Hochberg correction.

### 3.3 Association of nPAD with FSIQ

Patients with quotient scores outside two SDs were excluded (N = 47) to reduce bias in the estimation. The nPAD-WM and nPAD-multimodal scores significantly explained the variance (estimated beta of nPAD-WM = −1.428, *p* = 0.040; estimated beta of nPAD-multimodal = −1.426, *p* = 0.037; adjusted through Benjamini-Hochberg correction) in the FSIQ after adjustment for age, sex, and education years, indicating that the nPAD-WM and nPAD-multimodal scores were significantly and negatively correlated with the FSIQ. In contrast, the nPAD-GM score and FSIQ were marginally associated (estimated beta of nPAD-GM = −1.106, *p* = 0.089).

### 3.4 Structural deviance in schizophrenia based on ROI-based normative models

We used the ROI-based normative models to transform the structural features into Z-scores and to quantify the alterations of brain regions and tracts in patients with schizophrenia. The regions with significant alterations were identified using a mass one-sample *t* test for each feature with multiple comparison corrections, as displayed in Figure 3. The findings indicated that the MD index revealed more alterations than did the GFA index, and the regional volume was affected more than the cortical thickness.

**Figure 3.**
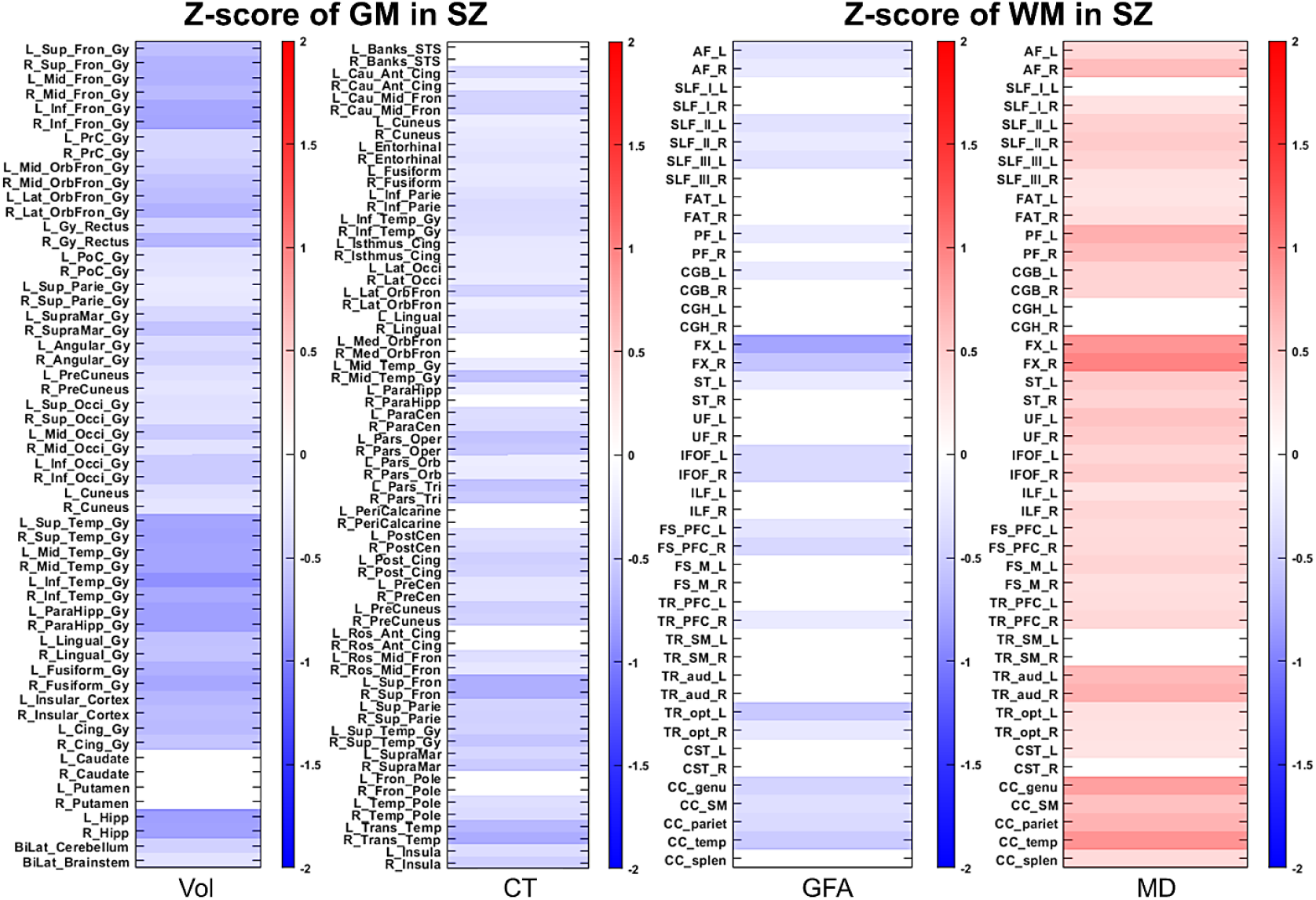
The density plots of the Z-score profiles in schizophrenia. The stripes with color coding indicate that the z-scores of the regions of interest in schizophrenia are significantly distinct from the norm whereas stripes with white color indicate no significant difference from the norm. The blue and red colors denote the negative and positive z-scores, respectively. Abbreviations: GM: gray matter; WM: white matter: SZ: schizophrenia; Vol: volume; CT: cortical thickness; GFA: generalized fractional anisotropy; MD: mean diffusivity. The full name of abbreviation is provided in Supplementary eTable.

### 3.5 Regional importance of apparent aging in schizophrenia

To investigate which underlying impaired regions contributed the most to the significantly increased nPAD scores in patients with schizophrenia, we calculated the coefficients using CCA (CCA in GM: rho = 0.995, CCA in WM: rho = 0.988) and normalized the values into a regularized range. The results are illustrated in Figure 4. Using descriptive statistics, we analyzed the importance of the top-half regions in each modality that were deemed to be representative of the most impaired regions and the greatest contributors to advanced aging in schizophrenia. The measurements of feature types in GM indicated that the regional volume (63.48% contribution rate) had a stronger effect than the cortical thickness (36.52%). In the WM measures, the MD values had a much higher rate of contribution (72.92%) than the GFA values did (27.08%). We further divided the GM features into five major anatomical regions: frontal, parietal, occipital, temporal, and limbic and other regions. The frontal (37.66%) and temporal lobes (35.86%) contributed the most, followed by the limbic and other regions (13.23%), the parietal lobe regions (9.18%), and the occipital lobe (4.08%). Features in the left GM hemispheric region (56.94%) contributed more to advanced aging than did features in the right GM hemispheric region (43.06%). In addition, the WM features were partitioned into the association, projection, and callosal fiber systems. The association system exhibited the greatest contribution (62.64%), followed by the projection fiber system (21.95%) and the callosal fiber system (15.42%). Furthermore, the WM features in the left hemisphere (48.41%) and right hemisphere (51.59%) exhibited relatively similar contributions.

**Figure 4.**
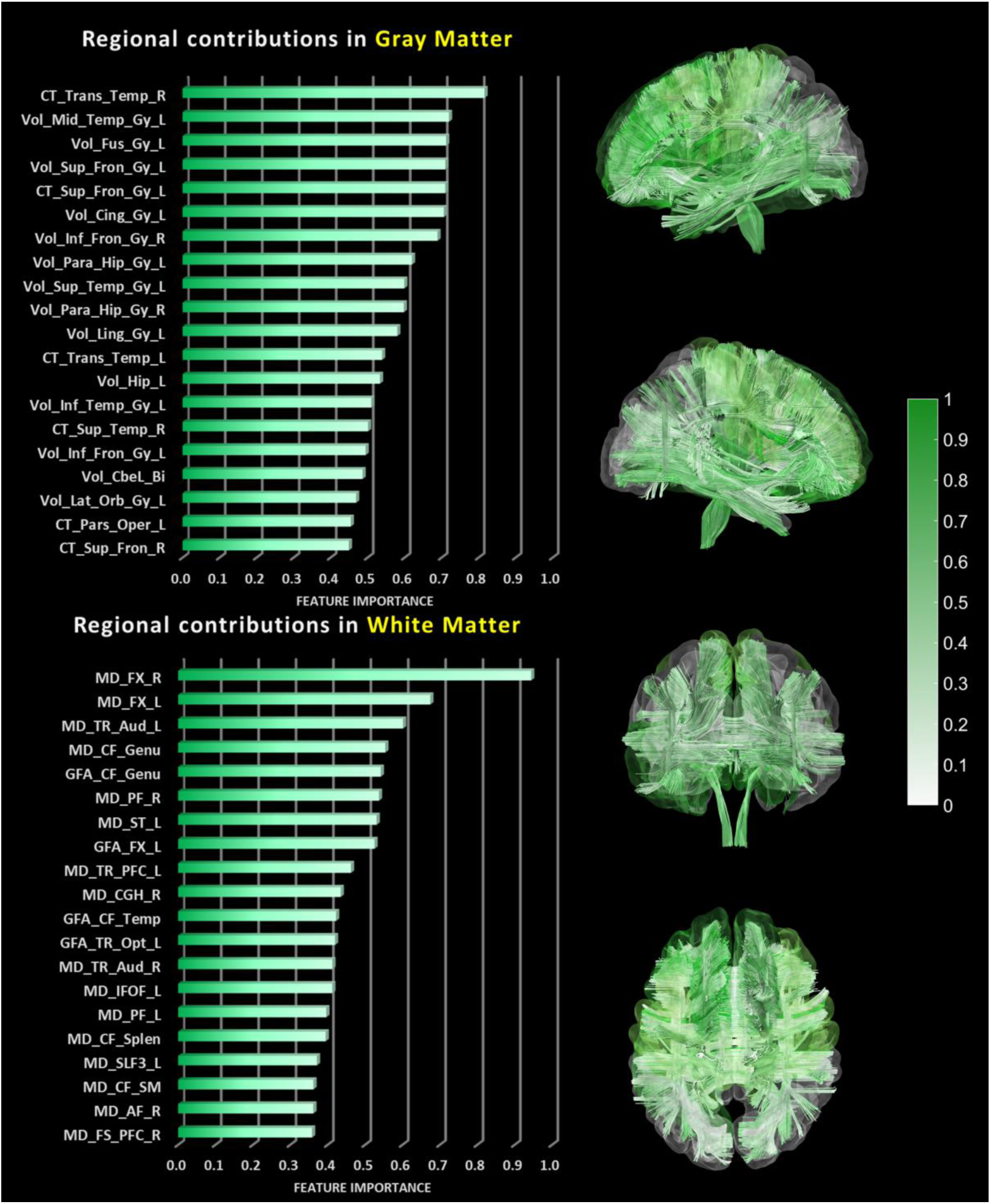
The bar graphs of regional importance in schizophrenia and the corresponding illustration in brain maps. The plots show the regions with the top 20 important features contributing to the normalized PAD scores of gray and white matters, respectively. The color spectrum encodes the importance of feature contribution. Abbreviations: CF: callosal fibers, CT: cortical thickness, Fus: fusiform, FX: fornix, Gy: gyrus, GFA: generalized fractional anisotropy, Inf: inferior, Lat: lateral, MD: mean diffusivity, Mid: middle, Par: para, Tem: temporal, TR: thalamic radiation, Vol: volume, Sup: superior.

## 4 Discussion

The etiology of schizophrenia has a substantive impact on aberrant maturational trajectories of the brain and subsequently leads to group-level differences in adult patients^3,45^. This study demonstrated that multifaceted brain age indices were capable of reflecting premature brain aging in schizophrenia, compared with the normal population. Particularly, the multimodal brain age index best distinguished the patients from normal brain aging, confirming that the multimodal brain age index had better sensitivity of reflecting the aberrant aging process^46^. Of the three models, WM brain age revealed significant associations with the age of disease onset and negative symptom scores, and the WM and multimodal brain age indices showed significantly negative associations with FSIQ. We also identified GM and WM regions in the frontal and temporal areas which contributed the most to premature aging, highlighting the neuroanatomical underpinnings of brain aging in schizophrenia.

The brain age measures of our study showed that premature brain aging in schizophrenia resulted from diffusively impaired brain structures over the whole brain. Our results of premature aging in GM replicate the previous findings, which reported that GM brain age was 3 to 5 years older than the normal^14,15,22^. This discrepancy is approximately equivalent to one standard deviation apart from the normal population based on the nPAD estimation. The aberrant aging in GM primarily reflects atrophic regional volume and reduced cortical thickness, and the former might be associated with premature aging, in line with the previous neuroanatomical observation^47^. Moreover, these findings are congruent with previous studies reporting widespread GM volume deficits and cortical thickness reductions in schizophrenia, most pronouncedly in the frontal lobe and temporal cortex^48,49^. Moreover, this pattern of neuroanatomical alterations in schizophrenia has been deemed to be similar to that in frontotemporal dementia (FTD)^50^. FTD refers to a neurodegenerative disorder which is characterized by the profound degeneration in frontal and temporal lobes, and primarily manifested in behavioral and personality abnormalities^51,52^. It has been reported that schizophrenia and FTD demonstrate certain commonalities in symptoms, etiology, genetics, epigenetics, and neuroanatomy^53,54^. Patients with FTD showed cortical atrophy and hypometabolism in frontal, temporal, cingulate, thalamic, and cerebellar regions^51-53^, and these abnormal regions were comparable to those observed in our GM regional contribution analysis. Additionally, the impaired structures in FTD appear to be lateralized; the left hemisphere is more severely impaired than the right hemisphere^51^. This also resembles our findings and previous reports in schizophrenia research^51,55^. Thus, we conjecture that the underlying mechanism of older brain age in schizophrenia might potentially have common causation with the process of neurodegeneration in FTD.

Likewise, the finding that the WM brain age was significantly older in schizophrenia is compatible with the study discovering the abnormal WM aging across the lifespan in schizophrenia^56^. Previous evidence has demonstrated that diffusion indices can reflect microstructural changes in WM and are sensitive to capture pathophysiological alterations in aging or mental disorders^56,57^. Through the use of brain age paradigm, we found that patients with schizophrenia showed group-level deviance of older-appearing brains in WM compared to their cognitively normal peers. Particularly, MD, which was deemed as a sensitive biological marker of disease and genetic liability in schizophrenia^58^, dominantly contributed to the observed premature aging. The MD increase might be related to tissue atrophy or fiber density reduction which might be related to pathological processes in schizophrenia^58,59^. Our findings also demonstrated that the association fiber system, especially those connecting to the frontal and limbic areas, played a key role in abnormal aging of WM in schizophrenia^60^. The impaired regions which dominantly contributed to the premature aging in schizophrenia might provide insight in treatment targeting. The WM abnormalities occur broadly in chronic patients, whereas the pathology might be confined to select fiber bundles which involve callosal fibers and those connecting to frontal and temporal areas early in the disorder^61,62^. In line with our findings, the fiber bundles connecting with the frontal, temporal, and limbic regions (e.g. fornix, genu, inferior fronto-occipital fasciculus, and callosal fibers connecting bilateral temporal lobes) were also substantially affected. Notably, most of these tracts functionally mature in early adulthood^63^, and this coincides with the time of peak risk for schizophrenia^61,64^. It has been hypothesized that developmental timing might confer increased susceptibility to disruption of particular tracts^61^. A stall in WM maturation may trigger psychosis^3^, and this would manifest as an onset-related decline in WM integrity that has been observed^65^. In our results, the WM brain age index was negatively correlated with the onset age, indicating that the earlier onset might be associated with the older brain age in WM, and this might imply that the earlier the impact in the maturational brain trajectory, the more severe the disruption of brain maintenance.

Although both GM and WM nPAD scores exhibited premature brain aging in patients with schizophrenia, the two scores merely showed a weak correlation. This implies that a universal premature aging process of GM and WM is ongoing in schizophrenia, but the senescence of these two structural metrics does not synchronize^66^. This is consistent with the finding that age-related abnormalities in GM and WM follow different temporal sequences in schizophrenia^67^. Another possibility of the weak relationship is that distinct clinical dimensions in schizophrenia might be related to aberrant developmental and aging trajectories in specific brain tissues.

From the perspective of symptomatology, different symptom dimensions of schizophrenia might have different neuroanatomical underpinnings, manifesting selective impairments in different structural dimensions. For instance, one study reported that associations of positive and negative symptom scores were found in WM but not in GM^68^. We also found that only the WM brain age measure was correlated with the negative symptom. From the perspective of cognitive impairment, it was reported that prefrontal WM but not prefrontal GM was correlated with various aspects of intelligence, including general abilities and working memory^69^. Similarly, we observed a significant negative association of the WM brain age index, but not the GM brain age index, with the FSIQ, suggesting that only older WM brain age in schizophrenia might prominently connect to more cognitive deficits. Therefore, different symptom dimensions in schizophrenia might be associated with distinct neural correlates, potentially contributing to different aging trajectories in GM and WM.

The illness onset and progression of schizophrenia have been conceptualized as a hypothesis called “early hit non-progression”^23,45^. In other words, the pathophysiological disruption of brain formation and reorganization occurs rapidly in the prodromal stage, and after the first episode, the progression of clinical syndromes is gradually maintained in a relatively stable phase called the “brain upkeep” phase in the chronic stage, which persists throughout the remainder of the lifespan^45^. Our negative finding on no associations between illness duration and brain age indices might be consistent with this hypothesis and the previous findings^21,23^. Although several studies claimed that the effect of aberrant brain aging became more prominent over time, especially within a short period after the disease onset^14,22^, we consider that, in a long-term scope, brain aging in schizophrenia might be relatively stationary in the remainder of the lifespan.

The findings of this study should be considered in light of several limitations. First, although we recruited a large number of patients with schizophrenia whose illness duration covered a wide range, the findings derived from the cross-sectional design are still limited to drawing inferences about individualized illness progression, which should be observed using a longitudinal design. Additionally, no demographically-matched controls were prepared for the case-control analysis. Nevertheless, a large number of cognitively normal subjects covering a broad life span suffice to offer a reliable reference for clinical samples. Finally, all of the patients used antipsychotic medication at the time of scanning. Although no correlation was found between antipsychotic medication and brain age indices, the medication effect might still be a potential confounding effect on brain aging regarding the nature of the illness. Future research is warranted to investigate the medication effect of brain aging in schizophrenia.

In conclusion, we have demonstrated that multifaceted brain age indices, which serve as a neuroimaging phenotype to reflect an individual’s aging status, are capable of detecting premature brain aging in schizophrenia. The WM brain age index is associated with the negative symptom severity, and the WM and multimodal brain age indices are negatively correlated with the intelligence quotient. Moreover, brain age exhibited associations with the onset age but no associations with the duration of illness, which are congruent with the early hit non-progressive hypothesis. In addition, we have identified the neuroanatomical contributions of premature brain aging in schizophrenia. The contributions mainly constitute the GM regions and WM connections within the fronto-temporal circuit, which resembles those impaired in FTD. This study provides detailed investigations of brain aging in schizophrenia using bias-free multifaceted brain age indices. Our findings add new knowledge to premature aging in schizophrenia, and might aid future studies to develop imaging biomarkers for stratification and prognostication in patients with schizophrenia.

## Data Availability

The data acquired from the National Taiwan University Hospital (NTUH) are not available due to the confidentiality agreement of the NTUH Research Ethics Committee. Code available upon request from the corresponding author.

## 5 Acknowledgments

This research was supported by the National Health Research Institute (NHRI) Taiwan (grant: NHRI-EX109-10928NI) and the Taiwan Ministry of Science and Technology (106-2314-B-002-242-MY3). The authors are grateful to the participants for their participation, the research assistants for assistance with subject recruitment and administration, and the technologists for performing MRI scanning. We thank Wallace Academic Editing for assistance with English editing that greatly improved the manuscript.

## 6 Conflicts of Interest

The authors declare that they have no financial/non-financial and direct/potential conflict of interest.

## 7 Author’s Contribution

C.C., T.H. and W.T. conceived the study and were in charge of overall planning. T.H., C.L., H.H., Y.L., M.H., C.L., and Y.C. enrolled and assessed the participants. C.C., Y.T., L.Y. helped in data collection of MRI scans. C.C., Y.T., L.Y. and Y.H. performed the image analyses. C.C. designed the prediction models, and conducted the experiments, statistical analyses, and visualization of results. C.C., T.H., C.L., H.H. and W.T. contributed to the interpretation of the results. C.C., T.H. and W.T. developed the theoretical framework and wrote the manuscript. All authors discussed the results and commented on the manuscript.

## 8 Ethical Approval

All procedures performed in this study involving human participants from the National Taiwan University Hospital (NTUH) were in accordance with the ethical standards of the NTUH Research Ethics Committee (REC) and with the 1964 Helsinki declaration and its later amendments or comparable ethical standards. Informed consent in the study was obtained from all individual participants who were recruited in the NTUH.

## Supplementary Information

S1 Brain Age Estimation

S2 Image Data Processing

S3 Results of Brain Age Models and Normalized Predicted Age Difference Estimation

References

## S1 Brain Age Estimation

### S1.1 Participants

The brain age models were created by using the neuroimaging data from National Taiwan University Hospital (NTUH) database. This database contained a training set (N = 482) and a test set (N = 70), which were used to establish brain age models and evaluate model performance, respectively (training set: mean age = 36.9 years, max = 92, min = 14, female proportion = 53.1%; test set: mean age = 36.8 years, max = 83, min = 14, female proportion = 52.2%). The distributions of age and sex in the 2 sets were statistically identical. The participants of the 2 sets were cognitively normal and met the recruitment criteria, including a Mini-Mental State Examination score of 25 or above, no self-reported substance abuse, no brain injury and brain surgery, no current experience of serious health problems, and no history of neurological diseases or psychiatric disorders. Participants who did not meet the safety and health-related criteria for MRI scanning were excluded.

### S1.2 Establishment of Brain Age Prediction Models

The details of the MRI imaging parameters and image processing are described in S2. After conducting image processing, the input data for gray matter (GM)-based brain age modeling used the volume and cortical thickness features in regions of interest (ROIs), whereas that for white matter (WM)-based brain age modeling used the tract-specific features of generalized fractional anisotropy (GFA) and mean diffusivity (MD). Consequently, the neuroimaging features of the GM- and WM-based model input consisted of 124 and 90 features, respectively. The sex factor was also included as a predictor in the models. Twelve-layer feed-forward cascade neural network models, which provide an accurate prediction with flexible model architecture for transfer learning, were used to predict age (4). The cascade neural network is a feed-forward neural network involving connections from the input and every previous layer to the subsequent layer. This network is similar to a simplified fully connected version of a dense block in densely connected convolutional networks, which avoid the vanishing-gradient problem and strengthen feature propagation (5). The loss function of model optimization was specified as a mean square error function, which was optimized using a gradient descent algorithm with an adaptive learning rate and constant momentum. A 10-fold cross-validation procedure was adopted within the training set to estimate brain age model performance. Validation set performance was used to stop the model parameter updates. The training procedure was implemented using MATLAB R2019a (MathWorks Inc., Natick, MA, USA) with an NVIDIA GeForce RTX 2080Ti (NVIDIA Inc., Santa Clara, CA, USA) graphics processing unit for accelerated computing. The performance of the trained brain age models was tested by predicting the brain age of individuals in the test set. To quantify model performance, Pearson’s correlation coefficient and mean absolute error between the predicted age and chronological age were calculated. The results of brain age modeling are provided in Supplementary Information S3.

## S2 Image Data Processing

### S2.1 MRI Imaging Parameters

The neuroimaging data in the NTUH database were acquired using a 3T Siemens TIM Trio scanner with a 32-channel phased-array head coil, with the same imaging protocol used for all data collection. High-resolution T1-weighted imaging was performed using a 3D magnetization-prepared rapid gradient echo (3D-MPRAGE) sequence: repetition time/echo time (TR/TE) = 2000/3 ms, flip angle = 9°, field of view (FOV) = 256 × 192 × 208 mm^3^, and acquisition matrix = 256 × 192 × 208; this resulted in an isotropic spatial resolution of 1 mm^3^. The imaging protocol for diffusion-weighted images followed that designed for diffusion spectrum imaging (DSI). The DSI datasets were acquired using the diffusion pulsed-gradient spin-echo echo-planar imaging sequence with a twice-refocused balanced echo (1, 2): TR/TE = 9600/130 ms, slice thickness = 2.5 mm, acquisition matrix = 80 × 80, FOV = 200 × 200 mm^2^, and in-plane spatial resolution = 2.5 × 2.5 mm^2^. The diffusion-encoding acquisition scheme used in this dataset followed the DSI framework published previously (2), in which 102 diffusion-encoding gradients were applied corresponding to the Cartesian grids in the half sphere of the 3D diffusion-encoding space (*q*-space) within a radius of 3 units equivalent to b_max_ = 4000 s/mm^2^ (3). Because the *q*-space data were real and symmetrical around the origin, the acquired half-sphere data were projected to fill the other half of the sphere.

### S2.2 Image Quality Assurance

Before we performed data analysis, all T1-weighted images underwent quality assurance (QA) procedures which are included in the Computational Anatomy Toolbox 12 (CAT12; http://dbm.neuro.uni-jena.de/cat.html), a novel retrospective QA framework for empirical quantification of quality differences. Retrospective QA involved automatic evaluation of essential image qualities such as noise, inhomogeneity, and image resolution. These quality measures were scaled to a rating scale, and “good” image quality level was required. Additional visual inspection was conducted to examine whether artifacts, including severe motion and abnormal lesions, remained in the images. All diffusion datasets also underwent QA procedures, including examinations for the signal-to-noise ratio (SNR), degree of alignment between T1- and diffusion-weighted images, and the motion-induced signal dropout (6). The SNR was evaluated by calculating the mean signal of an object divided by the standard deviation (SD) of the background noise (7). In practice, the signal was determined using a central square of an image for each slice, and the noise was averaged from 4 corner regions. Diffusion datasets with an SNR higher than mean SNR minus 2.5 SDs at their site were included. The degree of within-subject alignment between T1- and diffusion-weighted images was evaluated by calculating the spatial correlation between the T1-weighted image–derived WM tissue probability map and the diffusion-weighted image–derived GFA map. Higher spatial correlation indicated greater spatial alignment between T1- and diffusion-weighted images. In addition, because of the relatively long scan time of DSI, in-scanner head motion would inevitably cause signal dropout in diffusion-weighted images, particularly in those with high *b* values. For this reason, all participants lay on the MRI table with the head packed with expandable foam cushions to restrict head movement.

All acquired DSI datasets (5,712 images per participant) were examined by comparing the signal in the central square of each image with the predicted signal attenuation. Signal deviation from the predicted distribution was considered signal loss. Data with more than 60 images of signal dropout per participant (1% of the total diffusion-weighted images) were discarded.

### S2.3 Image Feature Processing

In the image feature processing for GM, voxel-based morphometry and surface-based morphometry were used to analyze the 3D MPRAGE data. The image analyses were performed using an extension of the Statistical Parametric Mapping package (SPM12; Wellcome Department of Imaging Neuroscience, London, UK; www.fil.ion.ucl.ac.uk/spm) (8) called CAT12. For voxel-based morphometry analysis, the structural imaging data were preprocessed using the default settings of the CAT12 toolbox, including corrections for bias-field inhomogeneity and segmentation into GM, WM, and cerebrospinal fluid, followed by spatial normalization to the ICBM template in MNI space (voxel size: 1.5 × 1.5 × 1.5 mm^3^) with SHOOT registration (9). The LONI probabilistic brain atlas, containing 56 ROIs, was used as a reference of volumetric tissue compartments (10) to estimate the volume of each ROI. For surface-based morphometry analysis of cortical thickness, we applied the automated surface-preprocessing algorithms included in the CAT12 toolbox that enable the estimation of cortical thickness of the left and right hemispheres by using the projection-based thickness method (11). Here, cortical thickness was determined by estimating the WM distance based on tissue segmentation. We used WM distance and a derived neighbor relationship to project local maxima (which is equal to the cortical thickness) onto other GM voxels. This approach included partial volume correction and correction for sulcal blurring and sulcal asymmetries. The Desikan–Killiany cortical atlas containing 68 cortical ROIs was employed to sample cortical features (12). In this manner, 56 volumetric features and 68 cortical thickness features were obtained to estimate GM-based brain age.

In the image processing for WM, we used an in-house algorithm called tract-based automatic analysis (13). First, the diffusion indices, including GFA and MD, derived from the diffusion MRI dataset were computed using the regularization version of the framework of mean apparent propagator (MAP)-MRI (14, 15). The signal in 3D diffusion-encoding space was fitted with a series expansion of Hermite basis functions, which describe diffusion in various microstructural geometries (16). The zero-order term in the expansion series contained the diffusion tensor that characterizes the Gaussian displacement distribution. Higher-order terms in the expansion series were the orthogonal corrections to the Gaussian approximation, and these were used for reconstructing the average propagator. The MD in each voxel was determined by calculating the mean of the 3 eigenvalues of the diffusion tensor (17, 18). We quantified GFA as the SD of the orientation distribution function divided by the root mean square of the orientation distribution function (19). To extract effective features of WM, the diffusion indices were sampled according to the spatial coordinates of 45 predefined major fiber tract bundles over the whole brain, which were constructed in the DSI template NTU-DSI-122 (20) through deterministic streamline-based tractography with multiple ROIs defined in the automated anatomical labeling atlas (21). In practice, the sampling coordinates were transformed from NTU-DSI-122 into individual DSI datasets with the corresponding deformation maps. The deformation maps were obtained through 2-step registration, which included anatomical information provided by the T1-weighted images (22) and microstructural information provided by the DSI datasets (23). The sampling coordinates were aligned with the proceeding direction of each fiber tract bundle, and diffusion indices were sampled in the native space along the sampling coordinates normalized and divided into 100 steps. Having sampled the diffusion indices, we averaged the indices across 100 steps along each tract bundle. Finally, 45 GFA features and 45 MD features were obtained for estimating WM-based brain age.

## S3 Results of Brain Age Models and Normalized Predicted Age Difference Estimation

We performed 10-fold cross-validation on the training set and the brain age models showed a strong linear correlation and low MAE between chronological age and predicted age based on GM features (rho = 0.956, MAE = 4.34), WM features (rho = 0.944, MAE = 4.76), and multimodal features (rho = 0.963, MAE = 3.99). The models also accurately predicted the brain age in the independent test set on the basis of GM features (rho = 0.943, MAE = 4.69), WM features (rho = 0.967, MAE = 3.95), and multimodal features (rho = 0.969, MAE = 3.97). Supplementary Figure 1 displays scatter plots of the predicted brain age in both the training and test sets.

After the brain age models were developed, normalization of predicted age difference (PAD) was conducted to minimize age-related bias (Supplementary Table 1). The results indicated that the approach was effective. For instance, the original PAD derived from the multimodal brain age model had a significant negative correlation with chronological age in both the training set (rho = −0.287, *p* < 0.001) and test set (rho = −0.308, *p* = 0.009), as displayed in Figures 1G and 1H, repsectively. After the predicted age was transformed into the normalized PAD (nPAD), the index did not show significant statistical bias with respect to chronological age in the training and test sets (rho = 0.013, *p* = 0.781, and rho = 0.028, *p* = 0.819, respectively), suggesting that the brain age indices were free of bias and could be used for further analyses.

**Supplementary table 1.** The linear correlation of PAD and nPAD with chronological age in the training and test sets.

| Models | Sets | Correlation b/w PAD and age | Correlation b/w nPAD and age |
| --- | --- | --- | --- |
| GM-based | training set | $\rho = -0.342$<br>$p < 0.001$ | $\rho = 0.026$<br>$p = 0.575$ |
| | test set | $\rho = -0.365$<br>$p = 0.002$ | $\rho = -0.031$<br>$p = 0.800$ |
| WM-based | training set | $\rho = -0.358$<br>$p < 0.001$ | $\rho = -0.016$<br>$p = 0.726$ |
| | test set | $\rho = -0.397$<br>$p = 0.007$ | $\rho = 0.054$<br>$p = 0.656$ |
| Multimodal-based | training set | $\rho = -0.287$<br>$p < 0.001$ | $\rho = 0.013$<br>$p = 0.781$ |
| | test set | $\rho = -0.308$<br>$p = 0.009$ | $\rho = 0.028$<br>$p = 0.819$ |

**Supplementary Figure 1.**
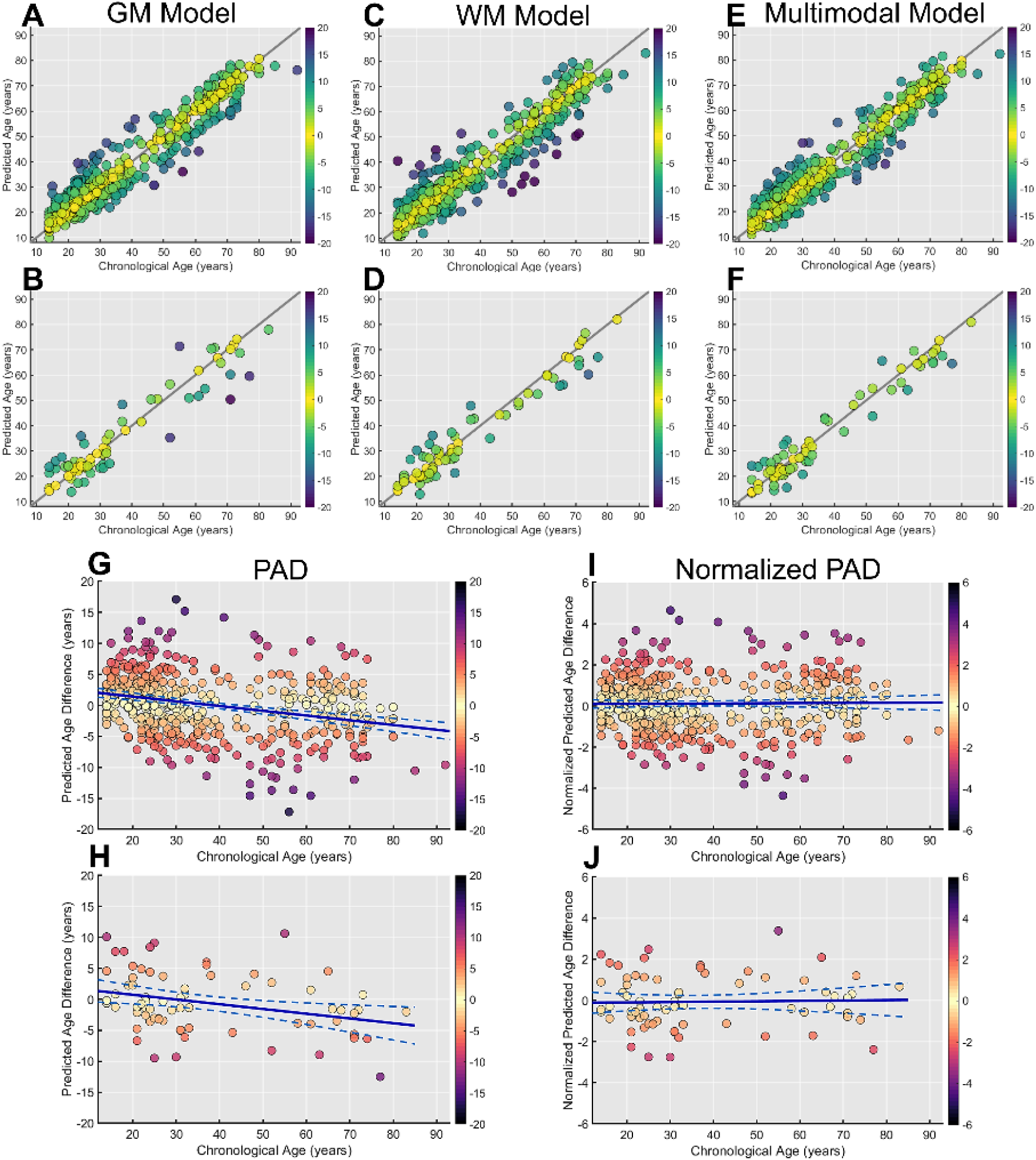
The scatter plots of chronological age against the predicted age derived from the brain age models in either the training set (A, C, E) or test set (B, D, F). The scatter plots of chronological age against the predicted age difference (PAD) are shown in the training set (G) and test set (H), and the scatter plots of chronological age against the normalized PAD are shown in the training set (I) and test set (J). Note that normalized PAD does not have age-related bias.

## Notes

### Competing Interest Statement

The authors have declared no competing interest.

